# Comparison of Antibody Levels in Response to SARS-CoV-2 Infection and Vaccination Type in a Midwestern Cohort

**DOI:** 10.1101/2021.08.16.21262036

**Authors:** Laura Remy, Chieri Tomomori-Sato, Juliana Conkright-Fincham, Leanne M. Wiedemann, Joan W. Conaway, Jay R. Unruh

**Author notes:** Correspondence to: Jay Unruh.

## Abstract

We present preliminary data in an ongoing observational study reporting SARS-CoV-2 spike protein reactive antibody levels from a convenience cohort of over 250 individuals in Kansas City. We observe stable antibody levels over one year in individuals who recovered from COVID19 infection caused by SARS-CoV-2. By comparison, our data reveals even higher antibody levels from naïve individuals vaccinated with Pfizer or Moderna vaccines and slightly lower levels from Johnson & Johnson (J&J) recipients. For all vaccines, inoculation after recovery resulted in higher antibody levels than vaccination alone. Responses to Pfizer and Moderna vaccines decreased over time from high initial levels but at the time of publication remain higher than those for recovered or J&J recipients. Within our limited cohort we only see slight demographic trends including higher antibody levels in recovered female vs. male individuals. Booster doses and breakthrough infections both result in rapid increases in antibody levels.

## Introduction

### Background/Rationale

The SARS-CoV-2 epidemic in the United States has stretched the limits of health care informatics, demanding real-time in-depth information about viral load and antibody responses across the country. This challenge was felt especially in the American heartland, where access to accurate and timely testing had been limited. Testing was often performed in response to suspected infections, rather than proactively across time, and antibody testing was rarely routinely performed. We present data from a convenience sample in the Midwest over 14 months. Subjects were routinely polymerase chain reaction (PCR) tested for SARS-CoV-2 and tested over time for antibodies to the SARS-CoV-2 spike protein.

### Objectives

Our objectives were to monitor levels of anti-SARS-CoV-2 Spike IgG antibody monthly in serum samples from a population with access to routine testing and to correlate those levels with infection, vaccination, and, where possible, demographic information.

### Setting

The Stowers Institute for Medical Research is a non-profit biomedical research institute located in Kansas City, Missouri. Stowers has approximately 550 employees including graduate students, post-doctoral trainees and support staff. At the beginning of COVID lockdowns in 2020, a routine surveillance PCR SARS-CoV-2 testing program was initiated to ensure safe return to work. In August 2020, our team began regular monthly collection of serum plasma for antibody detection on a convenience sample of employees and their household members. Data collection is ongoing.

## Methods

### Study design

Titration based ELISA testing was performed on volunteered whole blood samples once per month for 14 months. The target of the ELISA test was a synthetic purified spike protein (original strain modified for stability) (Amanat et al., 2020; Robbiani et al., 2020; Wu et al., 2020), which was detected using HRP labeled anti-human IgG secondary antibodies (Conkright-Fincham et al., 2021; Stadlbauer et al., 2020; Stadlbauer et al., 2021). The test was initially validated on commercial negative, active and recovered samples (Conkright-Fincham et al., 2021). For each sample, raw ELISA signals were plotted as a function of dilution, and results were reported as area under the curve (AUC) as calculated from a best fit to the data, normalized to the AUC obtained with the same set of dilutions performed on a sample of 20 ng/µL of a commercial anti-SARS-CoV-2 antibody (AM001414, Active Motif), such that a signal level of 20 represents equal area under the curve to the commercial antibody. Such a signal approximates a logarithmic transform of the titer value (Conkright-Fincham et al., 2021).

### Sample collection

Participants self-collected 100-150 µl fingertip blood specimens in capillary blood collection containers and serum samples were pre-processed as described in Conkright-Fincham et al (2021).

### Bias

Biases include cohort educational, economical, and racial bias (academic research center) relative to our community. Please see limitations section for detailed discussion.

### Statistical Methods

All error bars shown are standard error of the mean. In cases with two groups, t-tests were used for significance testing. For other groups, ANOVA was used.

### Ethical approval

The study was approved by the MRIGlobal Institutional Review Board (registration IRB00000067). Written informed consent for study participation was obtained from all participants.

## Results

### Participants

In total, 307 individuals inquired about this study of which 303 were enrolled. Study enrollment was limited to institute members and immediate family members over the age of 18. A total of 41 participants withdrew from the study for various reasons, including sample collection discomfort. At the time of this publication, there were 262 participants enrolled.

At the time of publication, 1465 testable samples have been collected from 263 individuals. There is a significant sample rejection rate, typically due to insufficient volume or self-collection error, with 144 samples rejected so far. This includes 141 identifying as females and 122 as males. Of the individuals contributing samples and reporting vaccination status, 21 reported receiving the J&J vaccine, 79 reported Moderna, 153 reported Pfizer, and 9 reported not receiving a vaccine. 43 individuals reported a covid infection resulting in a positive PCR test prior to vaccination, with 3 of those individuals reporting asymptomatic infections. 4 individuals have reported PCR demonstrated breakthroughs and 24 individuals have reported booster vaccinations. Participants giving samples were grouped by age with numbers shown in Fig. S4. Racial self-identification of participants giving samples included 213 reporting as white and 31 reporting as Asian. 19 individuals identified with other groups, including those ethnically reporting as Hispanic, that did not meet the threshold for separate analysis.

### Impact of SARS-CoV-2 infection

Samples from unvaccinated individuals reporting positive SARS-CoV-2 PCR tests were compared to those from unvaccinated individuals not reporting positive results. Those results are shown in Fig. 1A. Recovered individual samples displayed a range of signal levels. Recovered individual samples showed consistent and significant increases over those not reporting infection, with an average signal level of 23 for recovered individual samples compared to 1.2 for those not reporting infection. A small number of samples showed positive antibody signals without a positive PCR test. Those samples were ruled to be a result of asymptomatic infections after investigating contacts and symptoms and are not shown in Fig. 1A. Positive values followed an approximately normal distribution with a minimum signal of 10.

**Figure 1.**
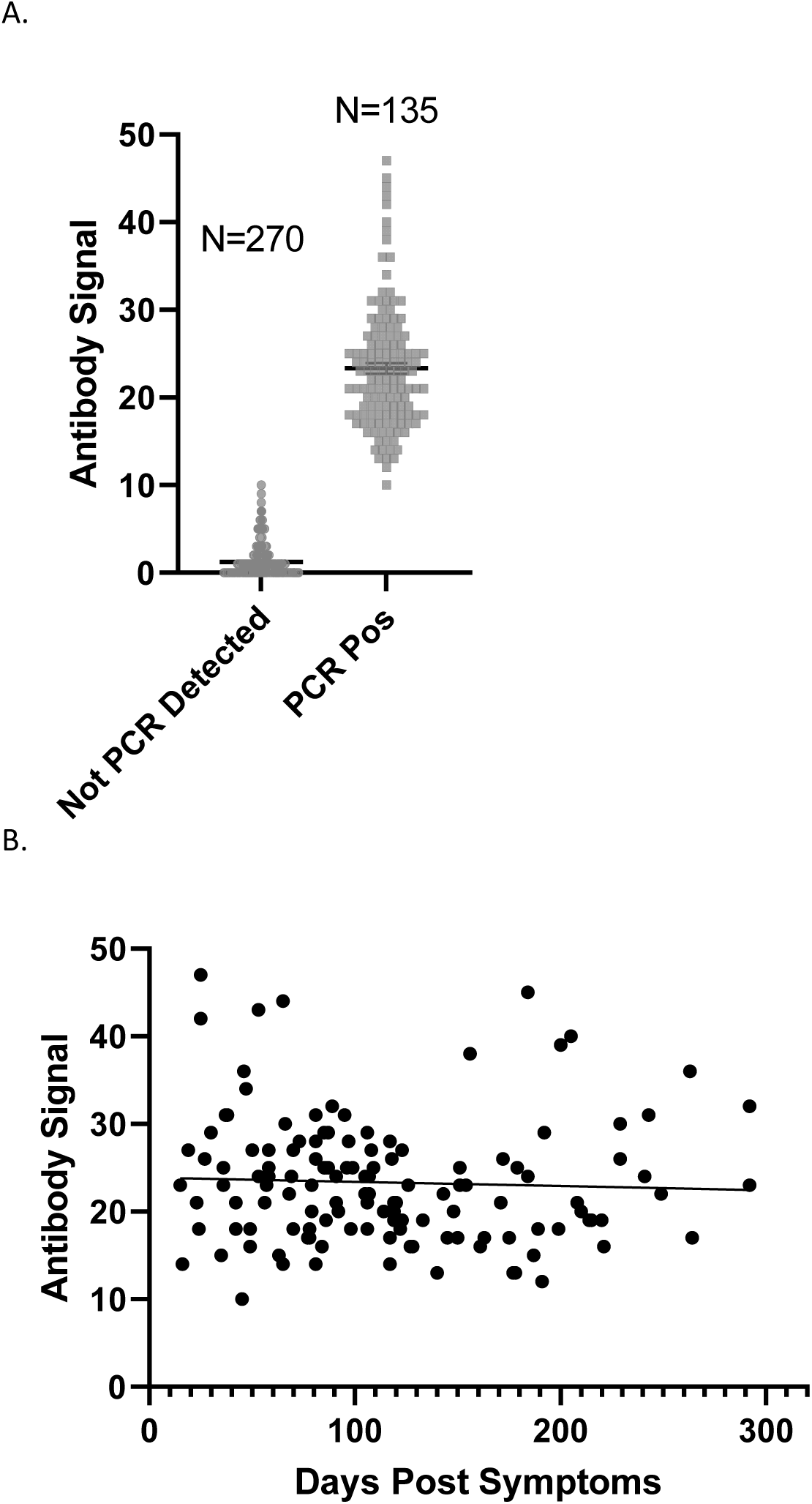
A.) Anti-SARS-CoV-2 spike antibody (IgG) signals (relative to AM001414) for unvaccinated individuals reporting positive PCR tests and those not reporting positive PCR tests. N refers to number of independently collected samples. B.) Antibody signals for PCR positive individuals from A as a function of time after symptom onset (or positive test if no symptoms were reported). The best fit trendline has a slope of -0.005 which is not significantly different than 0 (P = 0.61). Data include samples from 43 individuals.

### Impact of Time after SARS-CoV-2 Infection

Samples from unvaccinated individuals reporting positive SARS-CoV-2 PCR tests were recorded as a function of time after reported symptom onset. For asymptomatic individuals, the date of the positive test was used as the zero-time point. This data set includes multiple samples from many individuals. Fig. 1B shows those signals as a function of time. It is important to note that the assay detects IgG representing long lasting antibody signals. No significant decrease in antibody signals was detected from the aggregate data set over the time span measured during this study, though certain individuals may have shown a decrease. Three individuals were asymptomatic accounting for six of the samples included.

Previous studies have shown decreases in antibody levels over time (Long et al., 2020). It is possible that we do not observe this for our population because of the nature of our cohort or because of the nature of our assay. Certainly, limiting detection to IgG eliminates well documented decreases in other immunoglobulin species observed in some studies.

### Impact of Vaccination

Antibody signals as a function of time after reported vaccination are shown in Fig. 2A. Strong antibody responses are observed for Moderna and Pfizer vaccinated individuals starting around day 10 after the first dose of vaccine. After day 20, levels are variable but relatively stable with average signals slightly below 40. After 50 days, signal levels from Pfizer and Moderna begin to decline significantly with Moderna declining at a lower rate and showing significantly higher signals after 180 days. Data do not statistically support a non-linear fit model so we cannot predict if signals will level off in the future or continue to decline.

**Figure 2.**
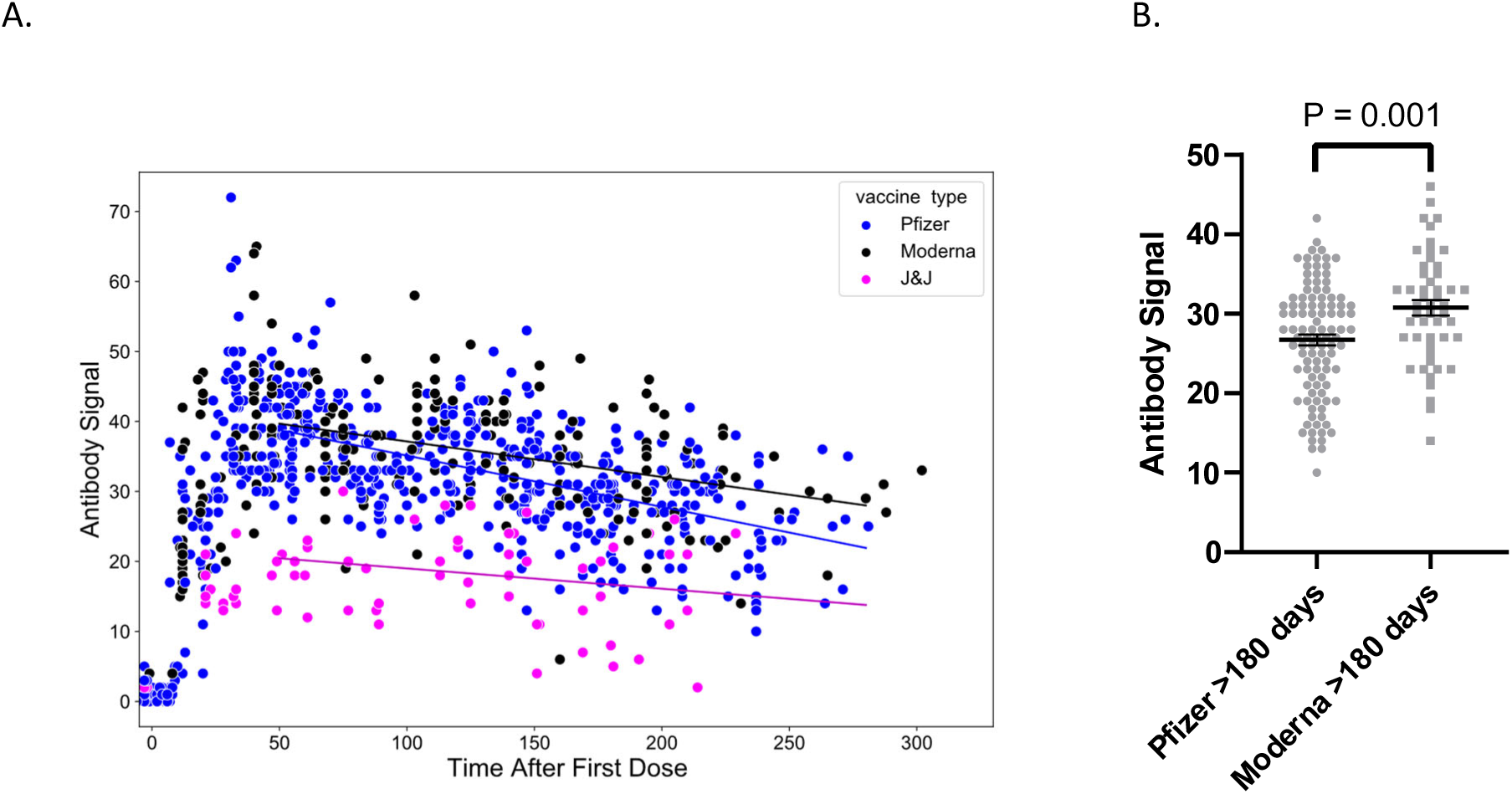
A.) Anti-SARS-CoV-2 spike antibody (IgG) signals as a function of time after vaccination for naïve individuals. Lines are best linear fit to data after 50 days. The J&J fit is not statistically different than slope = 0. The Pfizer and Moderna signals are decreasing at rates of 0.073 and 0.051 signal units per day, respectively, with P < 0.0001 for the slope differing from 0. No data set statistically supported a non-linear fit model. B.) Statistical comparison of Pfizer and Moderna signals more than 180 days after the first vaccine dose.

Fig. 3 shows statistical comparisons of different groups including recovered individuals more than 20 days after the first vaccine dose. J&J vaccine represents the lowest signal levels followed by recovered unvaccinated (slightly higher), followed by Pfizer and Moderna and recovered vaccinated groups. Combining Pfizer and Moderna signals results in a statistical increase for recovered vaccinated individuals above naïve vaccinated individuals with P < 0.0001. That result matches measurements from other studies as well (Bradley et al., 2021; Krammer et al., 2021). The timeline of vaccination response for recovered individuals does not show dramatic trends over time (Fig. S1).

**Figure 3.**
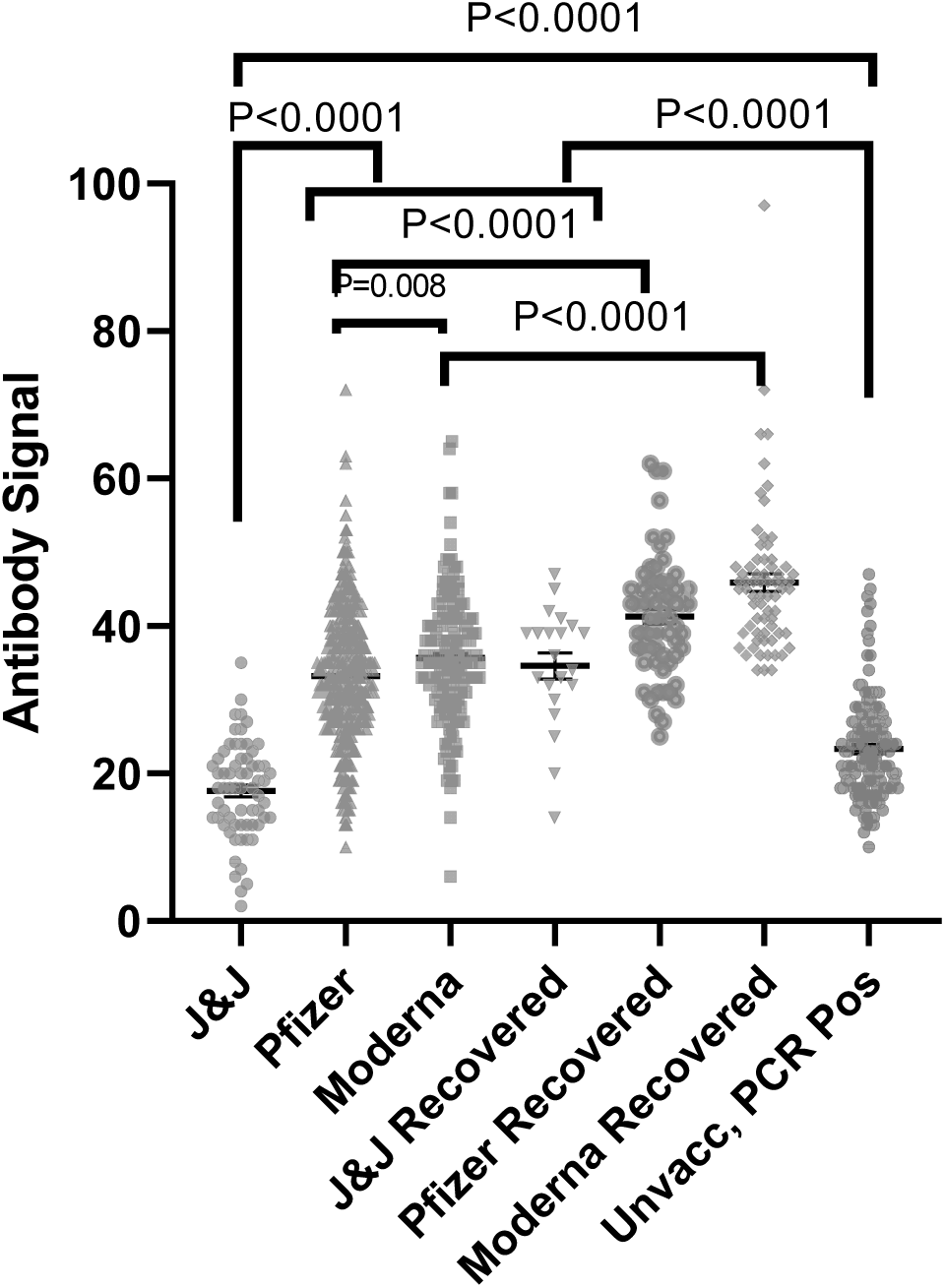
Anti-SARS-CoV-2 spike antibody (IgG) signals from samples from naïve and recovered individuals receiving vaccines from different manufacturers as compared to PCR positive unvaccinated individuals (data replicated from Fig 1). The numbers of independently collected replicates are (from left to right) 65, 197, 491, 21, 69, 63, and 135. Samples collected per individual are shown in Fig. S4.

These data clearly demonstrate the rapid increases in antibody binding in response to all vaccinations ten days after the first dose. The lower levels of antibody binding in response to J&J vaccination correlate with the lower reported efficacy of that vaccine. Nevertheless, is difficult to go beyond simple correlation given that our study measures binding only to one set of epitopes and does not measure neutralization or B cell production which are key factors in the immune response. At the time of publication, we do not have data on new variant binding which may differ from the binding of the original strain.

We have observed a small number of PCR confirmed breakthroughs all of which resulted in significant increases in antibody signals relative to previous vaccinated signals from the same individuals and from the entire cohort (Fig. 4A). Likewise, the individuals who have received Pfizer vaccinations and boosters experienced a significant increase in antibody signals (Figure 4B). These rapid and dramatic increases suggest that boosters are initiating a strong and rapid immune response.

**Figure 4.**
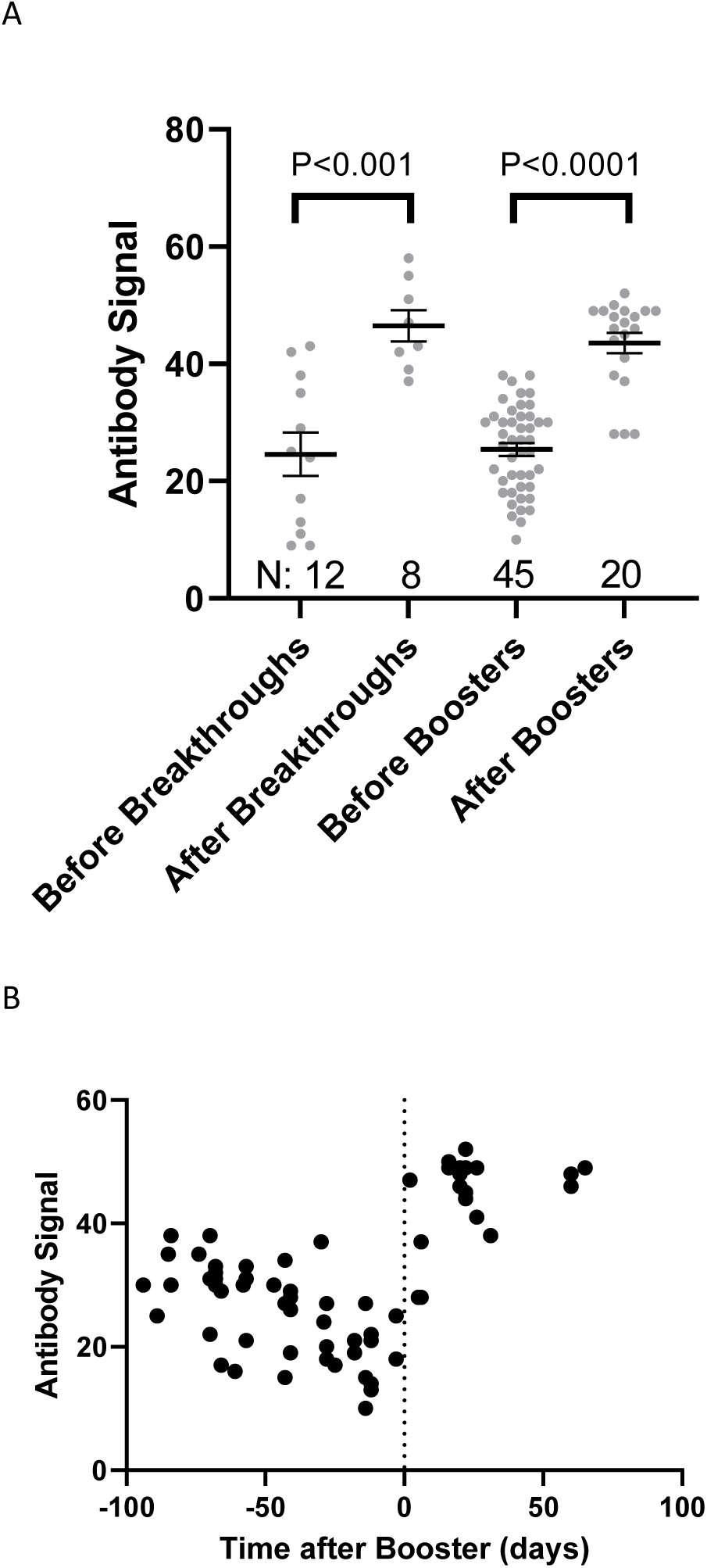
A.) Breakthrough infections and booster doses result in significantly increased Anti-SARS-CoV-2 spike (IgG) signal compared to the same individuals 100 days prior to those events. B) Time dependence of booster signal response. All booster signals were from Pfizer recipients.

### Demographic trends

We explored the demographic diversity of our data set to look for potential differences in antibody generation with gender, age, and race. Out of these demographic factors, only gender significantly impacted the antibody production in response to infection with SARS-CoV-2 (Fig. 5). While the male and female responses to Pfizer and Moderna vaccination were significant (P = 0.04) the magnitude of the response is small relative to the variation among individuals. We did not have enough data from individuals vaccinated with the J&J vaccine to compare gender response. We also did not have enough samples from recovered, unvaccinated individuals to measure the impact of race, but vaccination appears to produce a small but significantly higher antibody signal in individuals identifying as Asian (Fig. S2A). Increasing age decreases antibody signal slightly in response to vaccination as revealed by linear regression (Fig. S2B). Infection response does not appear to follow that same trend (Fig. S3). All of these factors cause far smaller differences than those between J&J/recovered and Pfizer/Moderna responses.

**Figure 5.**
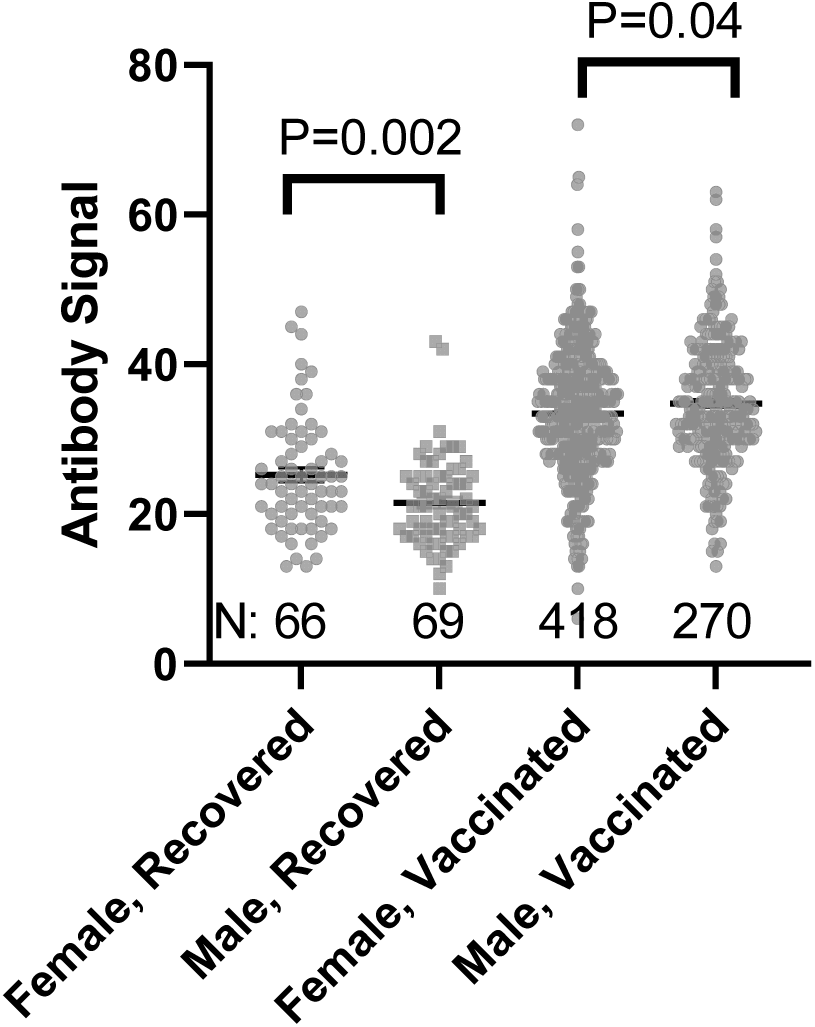
Comparison of Female and Male anti-SARS-CoV-2 spike antibody (IgG) signals. N refers to the number of independent replicates. Vaccinated group is only Pfizer and Moderna as other manufacturer groups did not achieve the numbers necessary for statistical comparison.

## Discussion

### Key Results

We have measured IgG in human serum via binding to spike protein in an automated ELISA assay over time for a small cohort in Kansas City undergoing frequent PCR testing. Our cohort showed sustained antibody signals after infection, extending to ten months after infection. Responses to vaccines from Moderna, Pfizer, and J&J were clearly seen ten days after the first dose and more consistently after 20 days. Antibody signals in individuals who received the J&J vaccine were significantly lower than in those who received vaccines from the other manufacturers and in recovered individuals, correlating with the reduced efficacy reported for the J&J vaccine (Sadoff et al., 2021). After vaccination with Pfizer and Moderna vaccines, responses peaked around 40 days after the first dose and trended downwards with Pfizer declining faster than Moderna. Future testing is needed to determine whether antibody levels continue to decline or whether responses tend to remain at levels similar to those seen in recovered individuals. In agreement with previous studies, all vaccines elicited a clear added increase in signal for those vaccinated after recovery with the most dramatic effect observed for J&J recipients. Likewise, breakthrough infections and booster doses rapidly increased in antibody levels above those for initial vaccination alone.

We observed little in the way of demographic impacts on antibody production, with only gender clearly impacting infection recovery. Antibody signal after Pfizer and Moderna vaccination decreased slightly with age and was higher in Asian individuals relative to their White counterparts.

### Limitations

Perhaps the biggest limitation of our data set is the focus on IgG binding to a single spike isoform. Variations in the spike protein as well as variations in the epitopes presented to serum antibodies are known to cause significant changes in antibody binding. In addition, IgA and IgM antibodies represent a strong initial antibody response to vaccination and infection and are well known to decay following response. Those limitations likely explain our lack of decay after recovery as reported elsewhere. Our assay also does not measure immune cell populations or viral neutralization, which are well known components of the immune response. Therefore, our antibody measurement represents a biased indicator of immune response over time and should not be interpreted on its own.

We also rely on self-reported vaccine data. Our data set suggests that that data is fairly accurate, but when considering outliers, reporting inaccuracies could have a significant impact.

Another limitation is uncertainty in collection method and timing. Samples were self-collected during a 24-hour period prior to serum preparation and freezing. While check-in steps should eliminate poorly collected and preserved samples, there is a possibility of contamination, timing error, and unknown collection factors that should be considered when interpreting the data.

Finally, it is important to note that our cohort, being volunteers from an academic research center, is a biased representation of our local community as a whole. As such, they are likely more educated and financially stable than our surrounding community. Racially, our cohort is more dominated by Asian and White participants than the general population. These factors dramatically affect access to and knowledge of health care and thus, our findings should be interpreted with those factors in mind.

### Data Availability

Original data underlying this manuscript is available from the Stowers Original Data Repository upon written request.

## Supporting information

Supplementary protocol manuscript draft

## Data Availability

Data is available on written request.

## Acknowledgements

We thank John Pak, Ph.D from Chan Zuckerberg Biohub for tips on spike protein expression and purification. We thank the Florian Krammer lab for the gift of the Spike protein expression vector. We are grateful to the Kansas City Community Blood Center for negative samples. We thank MIDC for positive serum samples used for initial validation of our assay. We thank the many individuals from Stowers who contributed to the assay development and execution to make this study possible and, in particular, discussions with Jennifer Gerton, Brent Kreider, and David Chao. A full list of those helping with the test is presented in the Supplemental Information.

## Author contributions

LR: study design, recruitment, data organization; C T-S: assay design and validation; J C-F: assay automation design, validation, and analysis; LMW: study design and biosafety assessment, JWC: study design, validation, and interpretation, JRU: principal investigator, study design, and data analysis.

