## Supplementary Figures and Text for "Comparison of Antibody Levels in Response to SARS-CoV-2 Infection and Vaccination Type in a Midwestern Cohort"

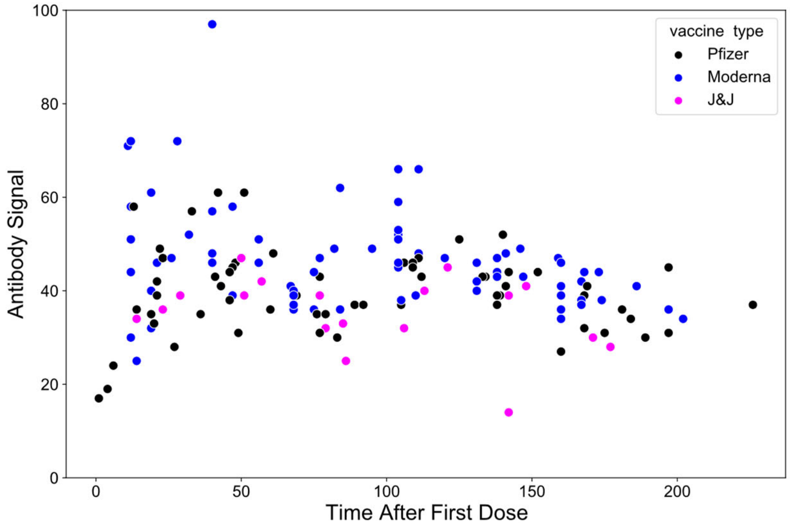


**Figure S1.** Anti-SARS-CoV-2 spike antibody (IgG) signal as a function of time for recovered individuals.

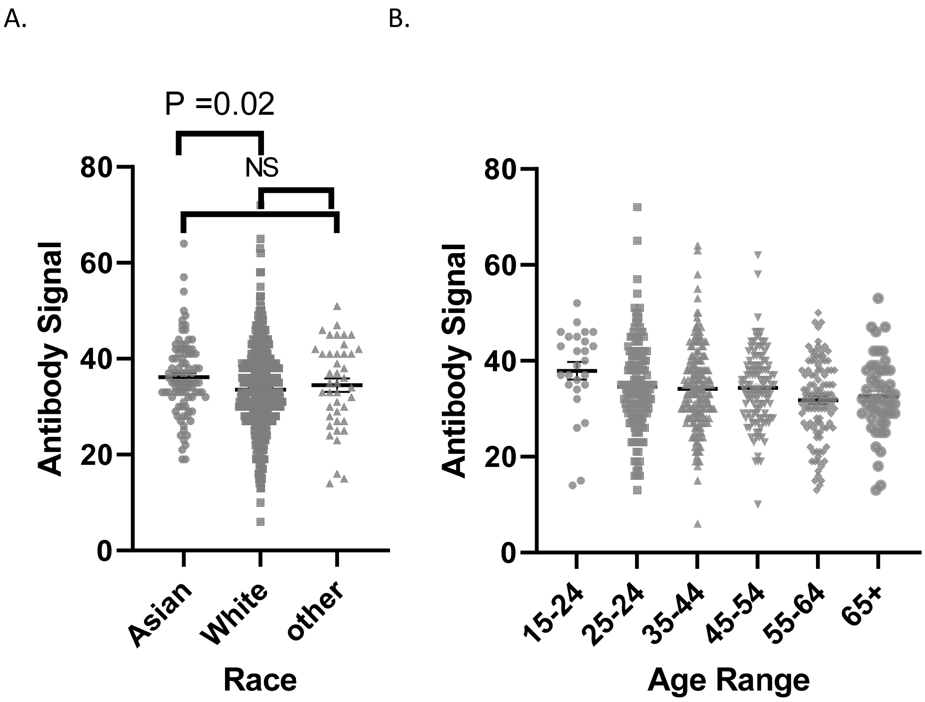


**Figure S2.** Anti-SARS-CoV-2 spike antibody (IgG) signal for individuals >20 days after first dose of Pfizer or Moderna as a function of individual race (A) and age (B). Participants identifying as Asian had a significantly higher antibody signal than those identifying as White. Regression of signal with age gives a significant downward trend with slope = -0.08 and $P < 0.001$.

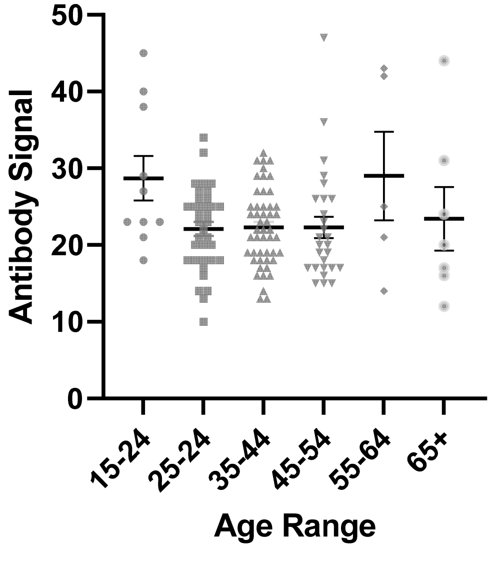


**Figure S3.** Anti-SARS-CoV-2 spike antibody (IgG) signal for recovered individuals as a function of age. ANOVA gives $P = 0.049$ for differences but there is no clear trend.

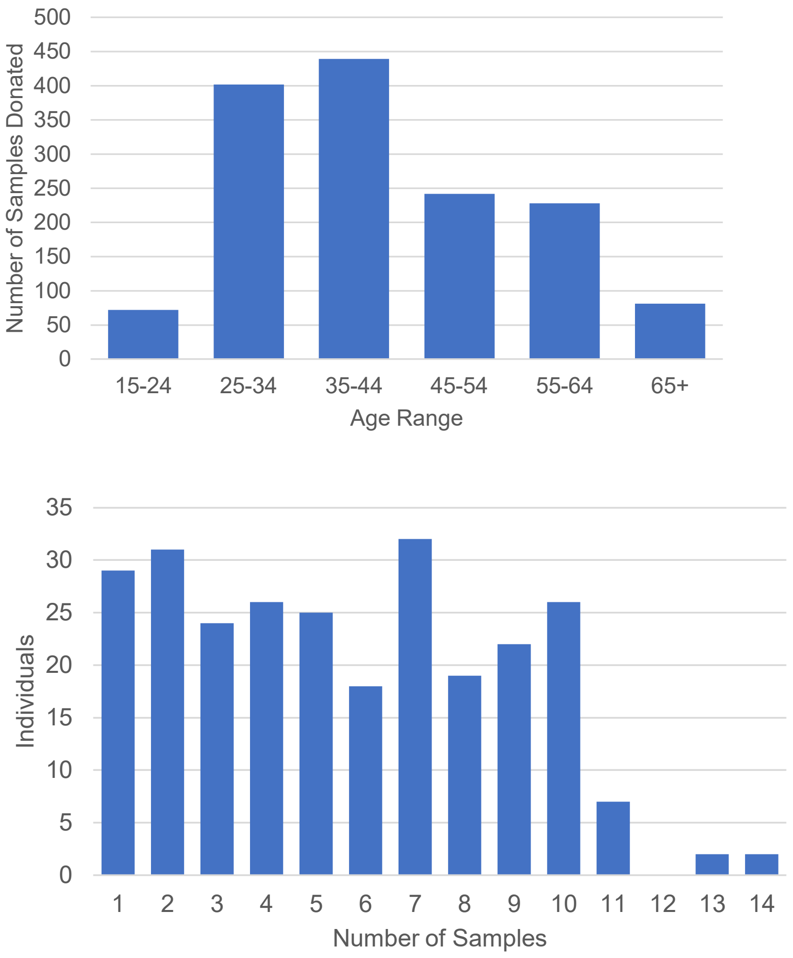


**Figure S4.** Numbers of participants giving samples as a function of reported age (top) and the distribution of individuals giving different numbers of samples (bottom).

**Supplementary Information:**

Additional people who are acknowledged for helping with the Stowers antibody study:

Adam Palmer, Alejandro Sánchez-Alvarado, Alexander Dean, Alexis Murray, Alice Hurley, Andrew Box, Andrew Hunn, Andrew Koebbe, Anoja Perera, Blake Bryant, Brandon Miller, Brandy Lewis, Brent Kreider, Brian Slaughter, Carolyn Beucher, Cindi Staber, Chongbei Zhao, Connie Hoye, Dan Bradford, Darren Wright, David Chao, David Karr, David Stiens, Denise Collins, Dorothy Stanley, Ed Weese, Ella Leslie, Gabe Keele, Heidi Olson, Jacob Yonke, James Summers, Jeff Haug, Jennifer Gerton, Jennifer Johnson, Jessica Witt, Jesus Gonzalez, Jim Selsor, John Bestor, John Kary, Judy Foye , Judy Zimmerman, Kelly Smith, Kevin Ferro, Kyle Weaver, Kym Delventhal, Lauren Weems, Laurie Ray, Leonardo Gomes de Lima, Lindsey Woolsey , Lisa Lassise, Madelyn Palmer, Maria Katt, Marshall Moore, Mary Penne Mays, MaryEllen Kirkman, Marysha Brown, Michelle Walker, Mike Newhouse, Nancy Thomas, Patsy Thompson, Pooja Chandra , Preeya Sharma, Rich McGee, Rhonda Wehrman, Robb Krumlauf, Robert Irby, Ron Conaway, Rory Fender , Russel Dorris , Sarah Rapp, Scott McCroskey, Stowers Security Team, Seth Malloy, Shannon Scott, Shigeo Sato, Shilpa Waduwawara , Stacey Walker, Tamara Potapova, Tara Gillett, Tari Parmely, Tim Geary, Tony Torrello, Tonyea Inglis, William Redwine, Yan Wang, Yongfu Wang, Xiangying Pi
