## Supplementary protocol manuscript draft for "Comparison of Antibody Levels in Response to SARS-CoV-2 Infection and Vaccination Type in a Midwestern Cohort"

### A High-throughput Automated ELISA Assay for Detection of IgG Antibodies to the SARS-CoV-2 Spike Protein

Juliana Conkright-Fincham[1]; Chieri Tomomori-Sato[1]; Rich McGhee[1]; Ella M. Leslie[1]; Carolyn J. Beucher[1]; Lauren E. Weems[1]; Shigeo Sato[1]; William B. Redwine[1]; Kyle J. Weaver[1]; Brandon D. Miller[1]; Kym M. Delventhal[1]; John J. Kary[3]; Andrew B. Koebbe[4]; Alexander Dean[1,5]; Jessica L. Witt[1]; Laura M. Remy[1]; Tari J. Parmely[1]; Chongbei Zhao[1]; Yan Wang[1]; Joan W. Conaway[2]; Jay R. Unruh[1*]

[1]Stowers Institute for Medical Research, Kansas City, Missouri, USA
[2]University of Texas, Southwestern, Dallas, Texas, USA
[3]Kary Software LLC, Lawrence, Kansas, USA
[4]Checkout 51 Inc, Toronto, Ontario, Canada
[5]Zan Consulting LLC, Erie, Colorado, USA

**[Abstract]**
The SARS-CoV-2 pandemic and vaccination campaign has illustrated the need for high throughput serological assays to quantitatively measure antibody levels. Here we present a protocol for a high-throughput colorimetric ELISA assay to detect IgG antibodies against the SARS-CoV-2 spike protein. The assay robustly distinguishes positive from negative samples, while controlling for potential non-specific binding from serum samples. To further eliminate background contributions, we demonstrate a computational pipeline for fitting ELISA titration curves, producing an extremely sensitive antibody signal metric for quantitative comparisons across samples and time.

**Keywords**: SARS-CoV-2, ELISA, Serology, COVID-19, Spike protein

**[Background]**
As SARS-CoV2 spread around the globe in 2020, the urgent need for assays to measure antibodies reactive to viral proteins became clear. Such assays are crucial as a first step for assessing immunological response to infection and vaccination. While it is clear that immunological responses are complex with many factors other than circulating antibodies contributing to protection after infection and long-term immunity, the ability to rapidly monitor antibody responses allows for population surveillance and provides the foundation for other, more complex, studies.

While several methods exist for colorimetric ELISA assays (Krammer and Simon, 2020), such methods are laborious, time consuming and require manual processing, which pose a safety risk to the researcher. We present a method based on previously developed ELISA titration protocols (Stadlbauer et al., 2020; Stadlbauer et al., 2021) a combination of semi-automated and fully automated steps for sample dilution and ELISA assay processes to allow for higher throughput in 384 well format with less human intervention. In addition, we present a computational method for fitting ELISA titration curves that accounts for varying background levels.

**Materials and Reagents**

1. PPE appropriate for working with SARS-CoV-2 materials
2. 3 MBS Capillary Tube (Micro Blood Science, Singapore)
3. Expi293F™ cells (Thermo Fisher Scientific)
4. Expi293 expression medium (Thermo Fisher Scientific, catalogue: A1435101)
5. Stericup 0.22 micron filter (Thermo Fisher Scientific, catalogue: SCGVU11RE)
6. HisTrap Excel™ column (Cytiva, catalogue: 29048586)
7. Phosphate buffered saline (PBS, standard recipe)
8. Protease inhibitor cocktail (Sigma, P8849)
9. 384w Immulon Maxisorp ELISA plates (ThermoFisher Scientific, catalogue number: 464718)
10. Universal plate lids (Fisher Scientific, catalogue 5500)
11. P30 384w tips for Janus MDT (PerkinElmer, catalogue number: 6001299)
12. 150ml automation reservoirs and base (Integra, catalogue number: 6318)
13. Bovine Serum Albumin (BSA) (SeraCare, catalogue number: 1900-0002)
14. Tween-20 (Sigma, catalogue number: P7949)
15. Plastic plate seal (Greiner, catalogue number: 676001)
16. MBS capillary blood collection containers (EDTA-2K #CH100ED2K) (Micro-Blood Science, catalogue number: 001001-001)
17. 0.3 ml Biobank tube in plate frame (Greiner, catalogue number: 9766561)
18. Axygen 96w U-bottom plate (Axygen, catalogue number: P-06-450R-C-S)
19. 125 µL Low retention Viaflo 96/384 XYZ GripTips (Integra, catalogue number: 6565)
20. Accurun 810 negative serum (SeraCare, catalogue 2010-0020) (stored at 4°C)
21. SARS-CoV-2 Spike Antibody, 414-1 (AM001414) (Active Motif, catalogue number 91361) (aliquoted and stored at -20°C)
22. 96w PCR plate (Axygen, catalogue number: PCR-96-FS-C)
23. Rack (plate frame) for 0.3 ml biobanking tubes (Greiner, catalogue number: 976501)
24. 125 µL Viaflo 96/384 XYZ GripTips (Integra, catalogue number: 6465)

25. Anti-human IgG (H+L)-HRP conjugated (Promega, catalogue number: W4031) (10 µL aliquots were stored at -20°C)
26. Normal goat serum (Jackson Immunoresearch, catalogue number: 005-000-121)
27. 1-Step Turbo TMB-ELISA substrate solution (ThermoFisher Scientific, catalogue number: 34022). Store at 4°C. Warm to ambient temperature prior to use.
28. Sulfuric Acid (JT Baker catalogue number: 9681-2)
29. PFM COVID-19 Active Symptomatic, Recovered (post-symptomatic) and Negative Serum (Precision for Medicine, special order). Aliquot and store at -80°C
30. Blood Center samples (for our work, this was kindly provided by the Kansas City Community Blood Center).

**Equipment**

1. AKTA Start (Cytiva) or equivalent FPLC system
2. Allegra X15X centrifuge with plate carrier rotor and carrier covers (Beckman Coulter)
3. Viaflo 96 base unit with 3 position stage and 5-125 µL 96w head (Integra)
4. Viaflo 96/384 base unit with 3 position stage and 5-125 µL 96w head (Integra)
5. LabElite decapper/scanner (Hamilton)
6. 405 TS washer/dispenser (405TSUVSQ) with 96 pin head (Biotek)
7. ELISA automated workstation contains the following components, which are integrated using a plate::handler II robotic arm (PerkinElmer) (F) and controlled with plate::works scheduling software (PerkinElmer).
    a. Stationary plate hotels (PerkinElmer)
    b. Automated 2x suction cup delidder (PerkinElmer)
    c. G3 Janus liquid handling platform with 4 PlateStaks with 4 magazines and 384w p30 MDT head (PerkinElmer)
    d. EL406 washer/dispenser with 192-pin head and peripump with 8-channel 10µL tubing (Biotek)
    e. Victor Nivo plate reader (PerkinElmer)

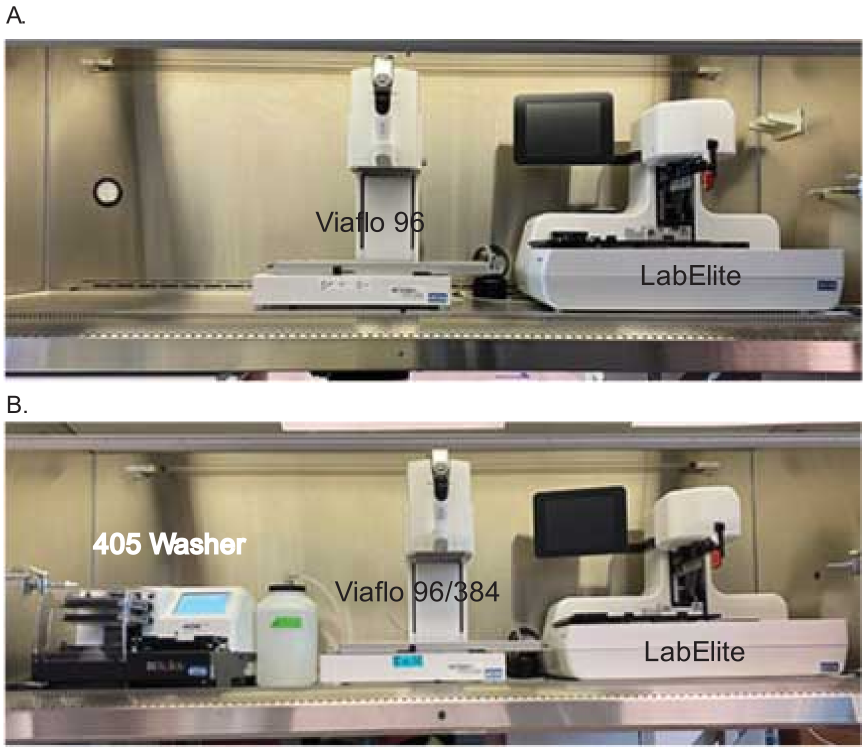


**Figure 1.** A) Sample dilution hood setup and B) ELISA hood setup.

A.

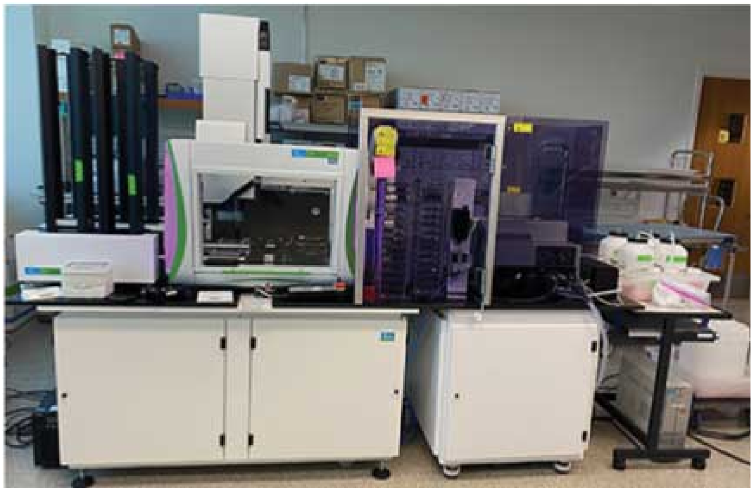

B.

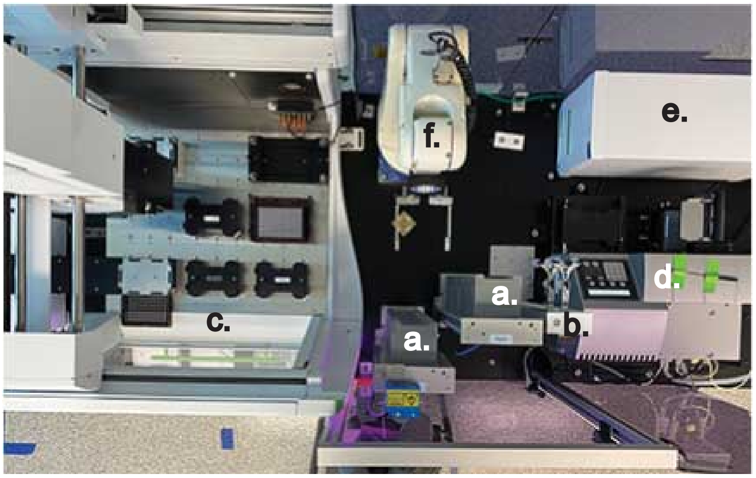


**Figure 2.** A) Side and B) top views of the ELISA automated workstation consisting of a) Plate hotels, b) suction cup delidder, c) G3 Janus liquid handler, d) washer/dispenser, e) Nivo plate reader, and f) robotic arm.

**Software:**

1. COVIDtrack custom sample tracking software (will be available on acceptance)
2. plate::works scheduling software (PerkinElmer)
3. Victor Nivo plate reader software (PerkinElmer)
4. Janus Winprep software (PerkinElmer)
5. Biotek LHC washer software
6. Microsoft Excel
7. Miniconda Python version 3 (https://docs.conda.io/en/latest/miniconda.html)
8. Custom python analysis code: https://github.com/jayunruh/Jay_Serology

**Procedure:**

A. Blood collection and plasma preparation

1. Participants in the study were assigned a unique QR code to be used for sample submission.
2. Blood collection kits included capillary blood collection containers labeled with a unique 1D barcode generated by COVIDTrack.
3. Participants self-collected 100-150 µL fingertip blood specimens in a barcoded capillary blood collection container and stored it at 4ºC for <24 hours before submission .
4. Submitted sample containers were inspected for leaks and required minimum volume (as indicated by marked lines on capillary) then wiped with ethanol.
5. Barcodes from each sample container were scanned and assigned to a Biobank tube and a 96-well position.
6. A 2D barcode on a 0.3 ml Biobank tube is scanned to assign the blood sample to a storage tube and placed in a matching well coordinate of a Biobank plate frame.
7. Sample containers were centrifuged at 2,600g for 5 minutes at 4ºC in a swing bucket rotor prior to carefully transferring 15-80 µL plasma to the assigned 0.3 ml Biobank tube and 96-well position.
8. The Biobank plate frame barcode is scanned, and the well location recorded.
9. Once all tubes have been scanned and designated as Accepted or Rejected, the scanned data is imported in COVIDTrack to update the location and status of samples.
10. Plasma was stored at 4ºC until assayed and then frozen at -70ºC for long-term storage.
11. To support deidentified sample processing, the Tube ID is converted to a Specimen ID following the sample check-in import. Well plate information including Specimen ID and well coordinates are downloaded from COVIDTrack for use in assay and data processing.

B. Spike protein production and purification

1. Plasmid containing the sequence for the modified spike protein was a gift from the Florian Krammer lab (Icahn School of Medicine at Mount Sinai, NYC). Modifications to remove furin cleavage sites and add cleavable purification tags (including 8x His) are as previously described (Amanat et al., 2020; Robbiani et al., 2020).
2. The insert was subcloned into an expression vector and stable Expi293F cell lines were established according to Tsai et al (2014).
3. Cells were cultured in suspension in Expi293 media on orbital shaker at 125 rpm in an 8% CO2 tissue culture incubator.
4. Clarified supernatants were obtained by centrifugation at 3000 rpm for 15 minutes followed by 0.22 micron filtration.
5. After adjusting pH to 7.4, cleared supernatant was applied to the HisTrap™ Excel column via the AKTA start system.

6. The column was washed with 50 column volumes of HisTrap wash buffer (containing 20 mM imidazole) supplemented with protease inhibitor cocktail.
7. Elution was performed with 10 column volumes of HisTrap elution buffer (contains 500 mM imidazole). Peak fractions were collected as a pool.
8. Buffer was exchanged to PBS with an Amicon Ultra column, yield was 1-2 mg per liter of culture.

C. Preparation of SARS-CoV-2 ELISA plates

1. The ELISA workstation was used to coat 384w ELISA plates with 20 µL of SARS-CoV2 Spike antigen diluted in PBS to a final concentration of 2 ug/mL in columns 3-12 and 15-24.
2. Columns 1, 2, 13 and 14 were coated with BSA by adding 20 µL of blocking buffer to these wells.
3. Plates were sealed with a plastic plate seal and placed at 4ºC overnight.
4. The following day, seals were removed from the coated plates and the plates returned to the workstation to be washed with wash buffer (4 x 80 uL).
5. Non-specific binding to the ELISA plates was blocked by adding 40 µL blocking solution to each well for 1 hour at room temperature and then washed with wash buffer (4 x 80 uL).
6. ELISA plates were used within 1 hour of washing.

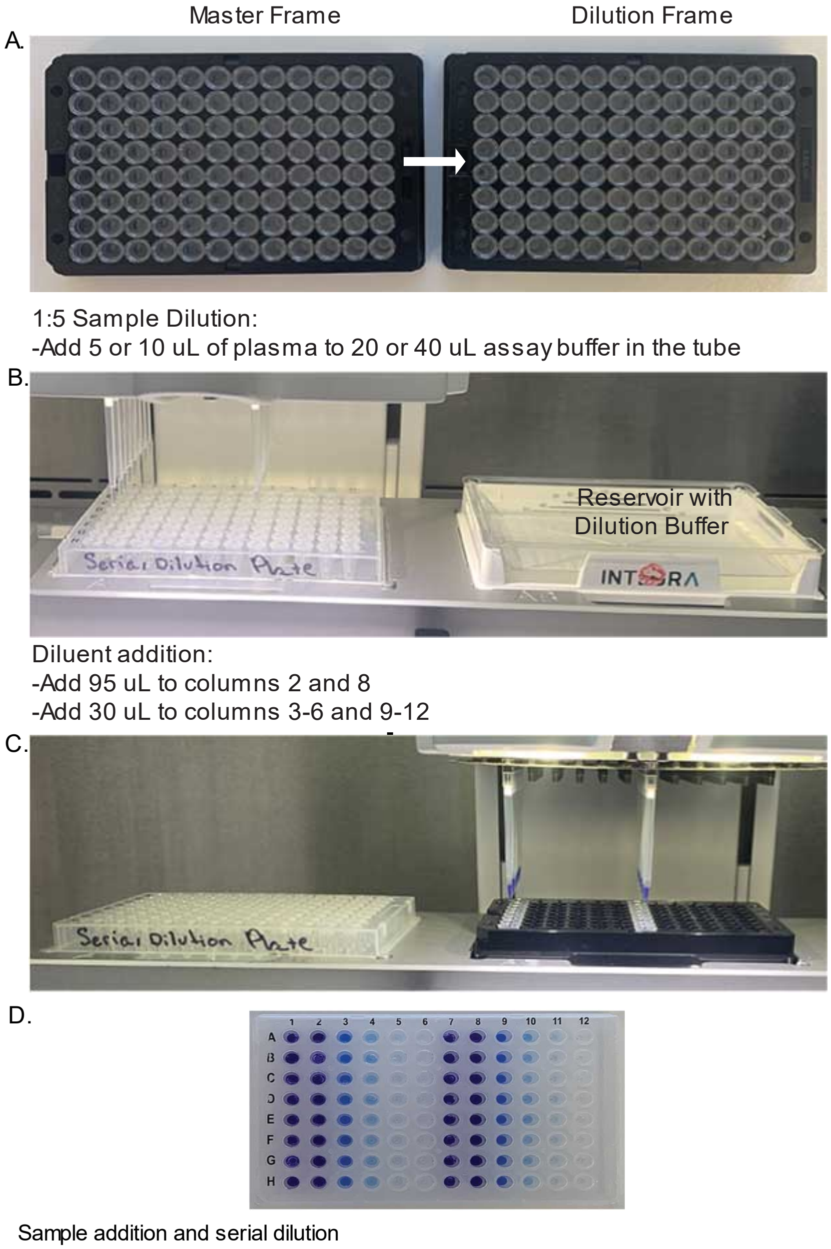


**Figure 3.** Flow chart for sample transfers and dilutions proceeding from top to bottom (A-D). In D, serial dilution occurs from left to right as described by procedure E and illustrated by blue dye.

D. Plasma Dilution (Fig. 3)

1. To collect the diluted samples at the bottom of the tubes, the biobank tubes in frames were centrifuged at 1000 RPM for 5 minutes at room temperature using a table centrifuge with plate carriers and locking lids.
2. In a biosafety cabinet, all samples (participant plasma, positive controls and negative controls) in 0.3 ml biobank tubes were decapped and were diluted 1:5 with plasma dilution buffer in separate 0.3 ml biobank tubes by using a Viaflo 96 well automated pipetting instrument with low retention tips. Tubes containing less than 20 μL volume were diluted manually.
3. Tubes were recapped using the decapper, and then centrifuged briefly at room temperature to ensure the diluted plasma was collected at the bottom of the tube.
4. Diluted plasma samples were then arrayed into columns 1 and 7 of each 96w source plate frame.

E. Serial Dilution (Fig. 3)

1. In a biosafety cabinet (dilution hood, Fig. 1A), the Viaflo 96 pipettor with low retention tips on columns 1 and 7 was used to dispense 30 μL of blocking buffer to columns 3, 4, 5, 6, 9, 10, 11 and 12 of a 96w PCR plate and 95 μL was added to columns 2 and 8 as the diluent for the serial dilution.
2. The 1:5 diluted plasma sample tubes were decapped and used as the source for the serial dilution. The Viaflo 96 pipettor was configured with low retention tips on the columns 1 and 7 to add 5 μL of diluted samples from the source plate frame into columns 2 and 8 of the PCR serial dilution plate (final conc 1:100) followed by slow mixing (3 x 80 uL).
3. 30 μL of the 1:100 dilution was transferred from columns 2 and 8 to columns 1 and 7. The source tubes were recapped and set aside, while the Viaflo 96 pipettor program continued to generate a 5-point 1:4 serial dilution by transferring 10 μL of diluted samples from columns 2 and 8 into columns 3 and 9 followed by mixing (3x30uL).
4. The process was repeated advancing the samples one column until 10 μL was dispensed into columns 6 and 12 and mixed.
5. 10 μL of diluted sample was removed from columns 6 and 12 and discarded. The final dilution concentrations in the series are 1:100, 1:400, 1:1600, 1:6400 and 1:25,600.
6. PCR plates were covered with lids and transferred to the ELISA hood (Fig. 1B).

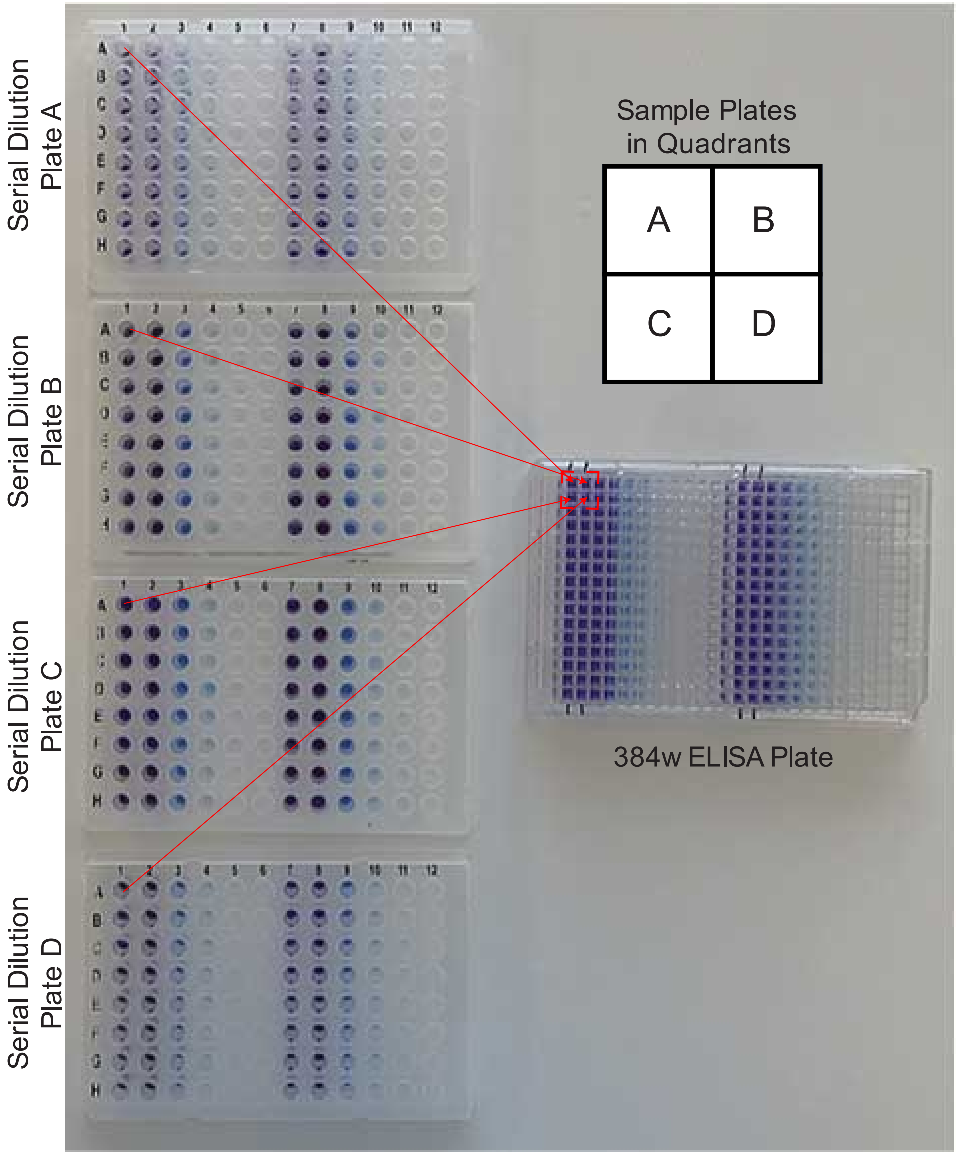


**Figure 4.** Addition of serial diluted plasma to quadrants of a 384 well ELISA plate as illustrated by blue dye.

F. ELISA 384 Well Assay

1. In a biosafety cabinet (Fig. 1B, ELISA hood), samples in 4-96w serial diluted PCR plates were assayed in each previously coated and blocked 384w ELISA plate by transferring 17.5 µL of each sample from the 96w serial dilution PCR plates into one of the four-well quadrants of the 384w ELISA plate (Fig. 4) using a Viaflo 96/384 automated pipettor with regular tips.
2. Inside the biosafety cabinet, ELISA plates with lids were incubated for 20 min at room temperature, and then washed (4 x 80 uL) with wash buffer.

3. After washing, the ELISA plates with lids were moved to the ELISA workstation, placed in the stationary plate hotels and the program executed.
4. Briefly, each plate in the run cycles through each of the steps in the ELISA workflow in a timed sequence.
5. The process started with the first delidded plate moving to the Janus deck and 20 µL of HRP solution was added to each well, then the plate was lidded and incubated for one hour at room temperature in the plate hotel.
6. Each plate was moved to the washer and washed (4 x 80 uL) with wash buffer.
7. To remove the residual Tween-20 in the wash buffer, 80 µL PBS was dispensed into the plate by the washer using the peri-pump followed by the washer removing the PBS from each well.
8. The robotic arm transferred the plate from the washer to the Janus deck to dispense 25 µL of Turbo TMB-ELISA substrate solution and returned the plate to the plate hotel for a 30 min incubation at room temperature.
9. The Janus system discarded the used tips into the tip box, which was upstacked into the PlateStak; while a fresh box of tips was downstacked to be loaded on the I30 MDT head.
10. To develop the assay, the plate was again moved to the Janus deck using the robotic arm and 25 µL stop solution was dispensed into each well.
11. Plates are transferred to the Victor Nivo reader and the absorbance of each sample read at 450 nm.
12. The robotic arm moved the plate to the washer to remove the stopped HRP reaction solution prior to returning the plate to the plate hotel for disposal.

G. Assay Validation

1. The 5-point ELISA assay was tested using a SARS-CoV-2 monoclonal antibody, AM1414, at increasing concentrations (2.5 ng/ml, 5 ng/ml, 10 ng/ml and 20 ng/ml) compared to the Accurun 810 negative serum (ARN) that the antibody was diluted in.
2. The AM1414 monoclonal antibody showed a dose dependent increase in signal (Figure 5A).
3. The Community Center Blood Center negative plasma (BB26, BB27 and BB29) had very little reactivity using the Spike antigen or BSA antigen (Figure 5B & C).
4. All PFM COVID-19 positive samples from active or recovered individuals show significantly higher absorbance values than the positive AM1414 control (Fig. 5D & E)
5. All PFM COVID-19 negative samples showed significantly lower absorbance than the positive AM1414 control (Fig. 5F) but higher absorbance values than blood bank samples (Fig. 5B).
6. Most samples showed no reactivity towards BSA coated plates, with the exception of PFM-R5 and PFM-N2 (Fig. 5G). Those samples were clearly distinguished as positive and negative, respectively, in the dilution assay indicating that non-specific reactivity is not a major limitation in our assay.

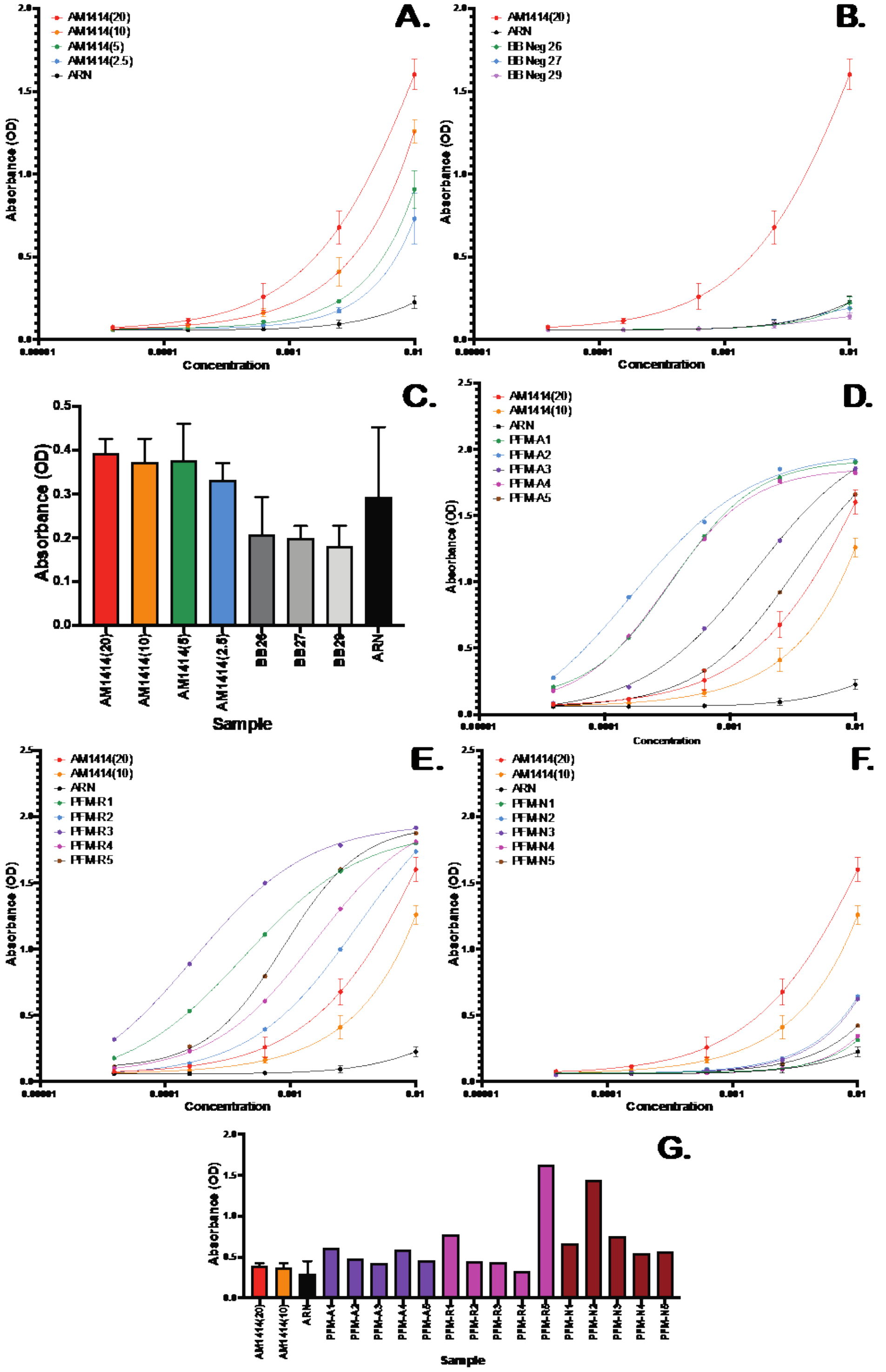


**Figure 5.** Validation of the Sars-CoV-2 Spike ELISA assay. Titration curves comparing A) different concentrations of AM1414 with the negative control (ARN), B) selected different blood bank samples, D) PFM samples from active infections, E) PFM samples from recovered individuals, and F) PFM samples from negative individuals. C and G show control, blood bank, and PFM control sample absorbance upon binding to wells coated in BSA.

H. Alternative 96 Well Procedure
1. The 384w assay was miniaturized from the initial 96w assay that was developed.
2. The ELISA workstation was configured for 96w assays by using the 96w I200 MDT head with P235 filtered disposable tips (PerkinElmer; catalogue number 6001289) on the Janus platform and the 96-pin head on the washer.
3. The basic workflow is the same; however, the serial dilution, volumes and timing is different.
4. The protocol is described briefly here. The 96w Immulon 2 HB ELISA plates (ThermoFisher Scientific, catalogue number 3455) were coated with 50 µL of Spike antigen in blocking buffer.
5. The coating was washed with wash buffer (3 x 250 uL) prior to the addition of 100 µL blocking buffer, which was washed with wash buffer after 1 hour incubation at room temperature.
6. The 1:5 plasma dilution was performed as described in the 384w assay.
7. The serial dilution was performed in low volume by adding diluent to the PCR plate (14 µL to columns 2 and 8 and 12 µL to columns 3-6 and 9-12).
8. 14 µL of plasma dilution buffer was added to columns 2 and 8 and mixed before 12 µL of the sample was transferred to columns 1 and 7.
9. A serial dilution was performed by transferring 4 µL of samples from columns 2 and 8 to columns 3 and 9, then mixing.
10. The process was repeated moving 4 µL of sample to complete the dilution, and subsequently, removing 4 µL from columns 6 and 12 to be discarded.
11. 108 µL of plasma dilution buffer was added to each well in the PCR plate and the plate transferred to the ELISA hood (Fig. 1B)
12. 100 µL of the diluted samples were transferred from the PCR plate to the coated and blocked ELISA plate and incubated for 1 hour at room temperature before washing (3 x 250 uL).
13. The ELISA plates were transferred to the ELISA workstation for the same workflow as the 384w plates with increased volumes of HRP solution (50 uL), wash buffer (3 x 250 uL), PBS rinse (250 uL), Turbo TMB-ELISA substrate (100 uL) and Stop solution (100 uL).
14. Plates were transferred to the Victor Nivo reader and the absorbance of each sample read at 450 nm.

#### Data Analysis

A. Titration absorbance analysis
1. Given the non-specific reactivity observed in our assay (Fig. 5) and the need for high throughput data analysis, we developed a custom ELISA analysis pipeline.
2. Analysis was performed in python with custom scripts (https://github.com/jayunruh/Jay_Serology). Raw absorbance values were organized by dilution concentration and fit to a simple binding model: $A(C) = b + A\frac{C}{EC50+C}$. This is equivalent to the Hill equation with a hill coefficient of 1.

3. Best fits were generated by searching over EC50 values starting at half of the lowest measured concentration and ending at 5 times the highest measured concentration. The search was performed nonlinearly with values multiplied by 1.05 at each step. At each step, A and b values were found by linear least squares and the minimum chi squared value from the search was used to identify the best fit values.
4. Binding was estimated using the area under the curve (AUC) defined as follows: $AUC = A * (C_{max} - EC50 * \ln(EC50 + C_{max}) + EC50 * \ln(EC50))$. This is simply the integral of the fit curve from 0 to the maximum concentration ($C_{max}$). Concentrations were defined relative to the starting 1/100 dilution.
5. Errors in individual fits were determined by the monte carlo method (Bevington and Robinson, 2003). Briefly, random Gaussian numbers were added to the best fit function with a standard deviation equivalent to the standard deviation of the fit residuals to simulate data with similar noise to the raw data. These simulations were then fit as described above. Errors are measured as the standard deviations of 50 simulations.
6. For every plate that was analyzed, we included three blood bank negative controls and a positive control of 20 ng/mL of AM1414 antibody.
7. We calculated a "Signal" measurement as follows: $Signal = 20 * \frac{AUC - Avg(AUC_{BB})}{AUC_{Pos} - Avg(AUC_{BB})}$. Therefore, the reported signal is 20 for a sample with the same AUC value as the positive control and 0 for a sample with the same AUC value as the blood bank samples. Signal values were rounded to the nearest integer and values below 0 were set to 0.
8. We observed that most experimental samples contained slightly more background than that observed in the blood bank samples. That background was matched by our purchased PFM negative samples, but we lack the quantities of those samples to use for everyday controls. Therefore, we estimated the statistics of the negative control by using the errors of the blood bank controls and the average values of the PFM samples adjusted up or down each day relative to the blood bank samples. Samples were called "non-negative" if they were different from the PFM mean values with a P value of <0.01.
9. Fig. 6 shows antibody signals from blood bank and PFM samples, validating our approach. Error bars are standard error in the mean.
10. The signal metric we measure has an approximately logarithmic relationship to the 1/EC50 value and therefore to the commonly measured titer value.

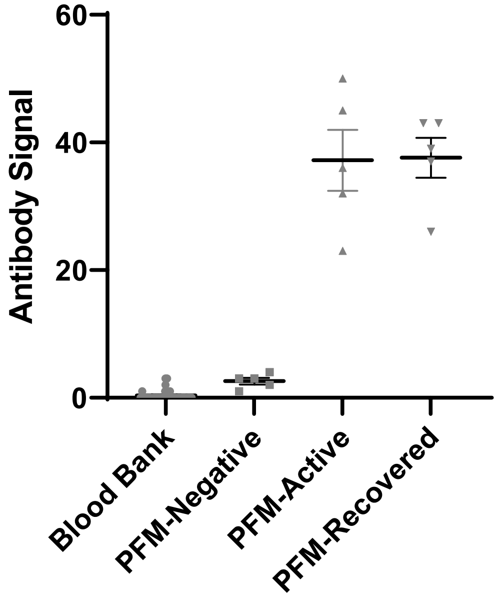


**Figure 6.** Quantification of commercial and blood bank samples demonstrating test accuracy.

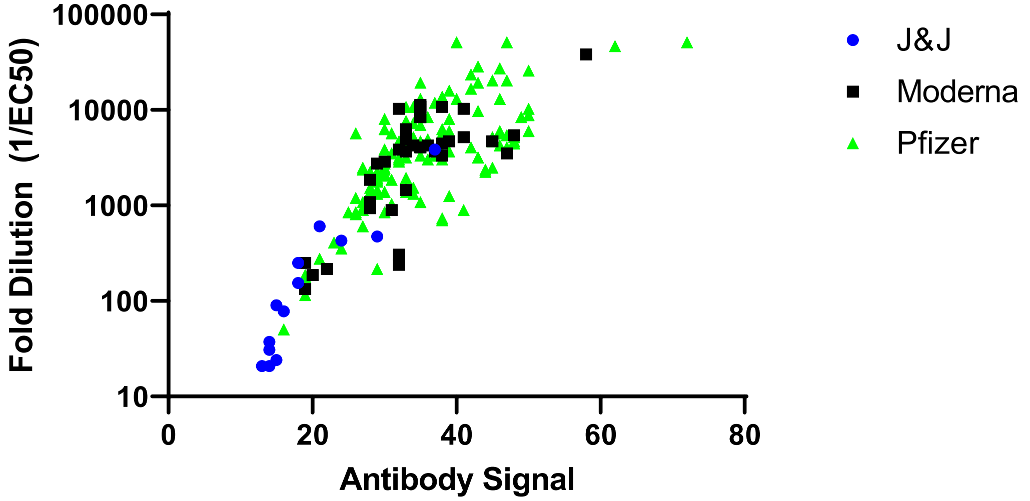


**Figure 7.** Comparison of EC50 and AUC values for vaccinated naïve samples over 20 days after the first dose. Other positive signals show a similar trend. While our method doesn't measure titer per se (such a value is noise dependent) the titer is approximately 11% of the EC50 value with a fold dilution 9 times higher than the value shown in this plot.

B. Result reporting
    1. Specimen ID and Biobank ID are assigned the test conclusion from the analysis (Positive, Non-Negative, and Negative) and a signal value.
    2. Data is imported into COVIDTrack to assign the results to the submitted sample and participant ID.
    3. Results and signal value are reported to participants via a confidential report.
    4. Data are organized by collection date and de-identified participant ID for downstream analysis.

**Notes**

1. Participant plasma was not heat inactivated; thus, potentially contains one or more pathogens and should be handled appropriately.
2. This assay can be performed with plasma or serum.
3. Plasma and serum are viscous; thus, slow aspiration and dispense speeds are required when pipetting them using automated instrumentation. Low retention tips are beneficial to prevent the plasma or serum from sticking to the plastic and reducing accuracy.
4. Hemolytic samples performed well in the assay.
5. Plasma or serum samples could be frozen and thawed at least 3 times without significant loss of signal.
6. Plasma or serum 1:5 diluted samples can be kept at 4°C for up to 3 days.
7. For BSL2 safety precautions, the ELISA plates were only processed on the workstation after the diluted plasma had been removed from the plate in the biosafety cabinet as it is open to the room air. In order to perform the entire ELISA on the workstation, the instrumentation would require an enclosure that filtered the air around the instrument through a HEPA filter to remove the possibility of aerosolization.

**Recipes**

1. HisTrap wash buffer consists of 20 mM imidazole in 20 mM sodium phosphate with 500 mM sodium chloride, pH 7.4.
2. HisTrap elution buffer consists of 500 mM imidazole in 20 mM sodium phosphate with 500 mM sodium chloride, pH 7.4.
3. 4X Plasma dilution buffer consists of 4% BSA diluted in 4X PBS pH 7.4, sterile filtered with a 0.45um Stericup (Sigma Millipore, catalogue number: S2HVU02RE) and stored in a 125 ml plastic bottle (Corning, catalogue number: 431731) at 4°C for at least 3 months. 1X Plasma dilution buffer was made by adding 75 mls of DI water to 25 mls of the 4X stock and then stored at 4°C
4. 4X Blocking buffer contains 4% BSA and 0.2% Tween-20 in 4X PBS pH 7.4, which was sterile filtered with a 0.45um Stericup and stored in a 125 ml plastic bottle at 4°C for at least 3 months. 1X Blocking buffer was made by adding 75 mls of DI water to 25 mls of the 4X stock and then stored at 4°C.
5. Wash buffer is comprised of 0.1% Tween-20 in 1XPBS pH 7.4.
6. AM1414 (20) positive control stocks were prepared by diluting AM001414 monoclonal antibody to 2000 ng/ml in Accurun 810 negative serum. 25 ml aliquots were made and frozen at -20°C for short-term storage or -80°C for long-term storage.

7. AM1414 (10) positive control stocks were prepared by diluting AM001414 monoclonal antibody to 1000 ng/ml in Accurun 810 negative serum. 25 ml aliquots were made and frozen at -20°C for short-term storage or -80°C for long-term storage.
8. AM1414 (2.5) positive control stocks were prepared by diluting AM001414 monoclonal antibody to 250 ng/ml in Accurun 810 negative serum. 25 ml aliquots were made and frozen at -20°C for short-term storage or -80°C for long-term storage.
9. 4X HRP buffer consists of 4% BSA and 0.4% Tween-20 in 4X PBS pH 7.4. 1X HRP buffer was made by adding 75 mls of DI water to 25 mls of the 4X stock and then stored at 4°C.
10. HRP solution was made by diluting human anti-IgG-HRP antibody 1:5000 in 1X HRP buffer containing 5% normal goat serum. This solution should be made immediately before use.
11. Stop solution was made by diluting concentrated sulfuric acid to 2M with molecular grade water.

**Acknowledgements**

We are grateful to John E. Pak from the CZI Biohub for advice on spike protein expression and purification. We are grateful to the Kansas City Community Blood Center for negative samples. We thank the Florian Krammer lab for the gift of the spike expression vector. We are grateful to MIDC for positive patient samples that were used to validate the assay but not included in this publication. Finally, we are grateful to the many Stowers Institute members who helped with this study, especially Jennifer Gerton, Brent Kreider, and David Chao for assay design discussions.

**Competing interests**

The author(s) declared no potential conflicts of interest with respect to the research, authorship, and/or publication of this article. While performing this research, no company had invested in the research, and no commercialization was intended.

**Ethics**

The study was approved by the MRIGlobal Institutional Review Board (registration IRB00000067). Written informed consent for study participation was obtained from all participants.

**Contributions**

J C-F: automation and assay development lead; C T-S: assay design and validation; RM, EML, CJB, LEW: assay automation design and validation; SS, WBR: spike protein purification and validation; KJW, BDM, and KMD: sample collection and check-in design; JK, AK, AD, JLW: sample tracking software development; LR: study design and sample tracking; TJP: study design and sample collection validation; CZ, YW: serum check-in processing and validation; JWC: assay design; JRU: data analysis and assay validation.

**Data Availability**

Commercial and blood bank data are available at https://www.stowers.org/research/publications/libpb-1656. Volunteer data is available from the Stowers original data repository on written request.

**Supplementary Information**

Movie: “Serology Assay-1.mov”: video of the robotic workstation running the assay.
